# Delay in Door-to-door-to-balloon time for Primary PCI is rarely Related to Cardiologists’ Late Arrival

**DOI:** 10.1101/2023.11.04.23298099

**Authors:** Mohammad Reza Movahed, Rana Irilouzadian

**Author notes:** Correspondence to: Mohammad Reza Movahed, MD, PhD, Clinical Professor of Medicine, Sarver Heart Center, 1501 N. Campbell Ave., Tucson, AZ 85724.

## Abstract

**Introduction:** Interventional cardiologists are held accountable for delay in the door-to-balloon time (DBT) for patients undergoing primary percutaneous coronary intervention in the setting of ST-elevation myocardial infarction (STEMI) even though in the chain of STEMI activation, the interventional cardiologist is the last person that needs to be available to start angiography. The goal of our study is to conduct a thorough analysis of the DBT data to assess time delays by randomly evaluating two consecutive years at the University of Arizona Medical Center (UAMC).

**METHODS:** We evaluated all available DBT data for STEMIs occurring in the fiscal years of 2011 and 2012 at the UAMC and calculated the time needed for the cardiologist to start the procedure after the patient was ready in the cardiac catheterization laboratory called Time to start the procedure (TSP) in addition to other time intervals.

**RESULTS:** Mean TSP time was 4 minutes and 24 seconds, one of the shortest time delays in the chain of STEMI activation and DBT. The median TSP delay was 3 minutes. The longest delay interval was the STEMI team’s arrival to with a mean of 17 minutes and 38 seconds.

**CONCLUSIONS:** Our data is the first to evaluate delays related to DBT revealing the least delay occurring due to the late arrival of Interventional cardiologists. Our data emphasizes the importance of performing a detailed time analysis of the DBT delay in order to objectively determine the actual areas of delay and provide a future pathway to improve them since we have specifically detected a delay in STEMI team and patient arrival to the catheterization laboratory as the main delay in the DBT time. in order to avoid blaming the wrong person and find the true root cause of the delay.

## Introduction

ST-segment elevation myocardial infarction (STEMI) is a life-threatening condition requiring reperfusion therapy as soon as possible (1) with primary percutaneous coronary intervention (PCI) to be performed if available in a timely manner. The time to reperfusion therapy encompasses two components: the interval from symptom onset to the initial call for assistance, and the system delay (the time from patient contact with the healthcare system to reperfusion). While organizational measures can enhance the system delay, patient delay is influenced by multiple factors. System delay serves as a prognostic indicator for mortality in STEMI patients undergoing primary PCI (1-3). So, one of the key factors in effectively managing patients with STEMI is minimizing the time from hospital arrival to balloon inflation, commonly referred to as door-to-balloon time (DBT). Longer delays in DBT have been associated with poorer clinical outcomes and increased mortality rates. Therefore, a primary objective in the management of such patients is to reduce DBT, as it plays a crucial role in mitigating in-hospital mortality risks (4).

Several studies have explored the relationship between DBT and the mortality rate among patients with STEMI. An established cardiovascular goal for such patients undergoing primary PCI is achieving a DBT of less than 90 minutes for at least 85% of the patients. Evidence indicates that briefer sessions of DBT, particularly those lasting less than 60 minutes, show a marked decrease in the likelihood of recurrent myocardial infarction within 30 days, along with lower rates of both in-hospital and 30-day mortality (3, 5-8). The interventional cardiologist, though the final person required for angiography initiation, is held responsible for delays in DBT. However, few studies have evaluated the specific components of DBT to determine which aspect plays the most prominent role within this time frame (9-11). Accordingly, we aimed to conduct a comprehensive analysis of the available DBT data for all STEMI cases that were referred to the University of Arizona Medical Center in a one-year period.

## Methods

This is a retrospective observational study that was conducted at the University of Arizona Medical Center (UAMC). We randomly evaluated two consecutive years for all available DBT data for STEMIs occurring in the fiscal years of 2011 and 2012. The study was approved by the Institutional Review Board. All clinically confirmed consecutive cases of acute STEMI occurring in these two study years were included in the study. Patient data were collected. The timetable of the steps involved in taking the patients from the place where STEMI was diagnosed until the procedure was started was examined.

Data of the following times were collected: Patient arrival to the emergency department (ED), obtaining ECG, STEMI read on the ECG, STEMI team arrival in the cardiac catheterization laboratory, patient arrival time in the cardiac catheterization laboratory, patient ready time which is the time that patient is prepped and draped and ready for the procedure to start, begin time which is the time when the physician has arrived, scrubbed and local anesthesia is injected at the procedure site and reperfusion time. Using the collected times, the following intervals were calculated: Patient arrival to ECG, ECG done to STEMI read, STEMI team activation to STEMI team arrival, STEMI team arrival to patient arrival in cath lab, patient arrival in cath lab to patient ready, patient ready to begin time (time to start procedure, TSP), begin time to reperfusion time, and DBT.

## Results

The data of 76 STEMI patients who underwent PCI was collected. The duration of all of the times of the patients is presented in Table 1. The mean duration of DBT was 68 minutes and 17 seconds. The longest delay was the time of the STEMI team’s arrival to the patient’s arrival in the cardiac catheterization laboratory with a mean of 17 minutes and 38 seconds and the median delay for this time was 16 minutes. Mean TSP time was 4 minutes and 24 seconds, one of the shortest time delays in the chain of STEMI activation and DBT. The median TSP delay was 3 minutes. The dot plots of all of the STEMI patients for DBT, patient arrival in procedure room to patient ready, patient ready to begin time, and begin time to reperfusion time are presented in Figures 1-4.

**Table 1.** Timetable for STEMI Patients Undergoing primary percutaneous coronary intervention.

| **Interval** | **Minimum** | **Maximum** | **Median** | **Mean** | **Std. Deviation** |
| --- | --- | --- | --- | --- | --- |
| **Patient Arrival to ECG obtain, min.** | 0.00 | 27.00 | 4.00 | 5.17 | 4.74 |
| **ECG done to STEMI read, min.** | 0.00 | 68.00 | 0.50 | 3.47 | 8.77 |
| **STEMI Team Activation to STEMI Team Arrival, min.** | 1.00 | 35.00 | 9.00 | 10.57 | 10.04 |
| **STEMI Team Arrival to Patient Arrival in Cath Lab, min.** | 0.00 | 46.00 | 16.00 | 17.63 | 10.61 |
| **Patient Arrival in Cath Lab to Patient Ready, min.** | 0.00 | 69.00 | 8.00 | 8.96 | 8.69 |
| **Patient Ready to Begin Time (TSP), min.** | 0.00 | 25.00 | 3.00 | 4.39 | 5.11 |
| **Begin Time to Reperfusion Time, min.** | 2.00 | 91.00 | 16.00 | 19.15 | 13.73 |
| **DBT, min.** | 22.00 | 196.00 | 68.00 | 68.28 | 24.34 |

Abbreviations: DBT, door-to-balloon time; ECG, electrocardiography; min., minutes; STEMI, ST elevation myocardial infarction; TSP, time to start the procedure

**Figure 1.**
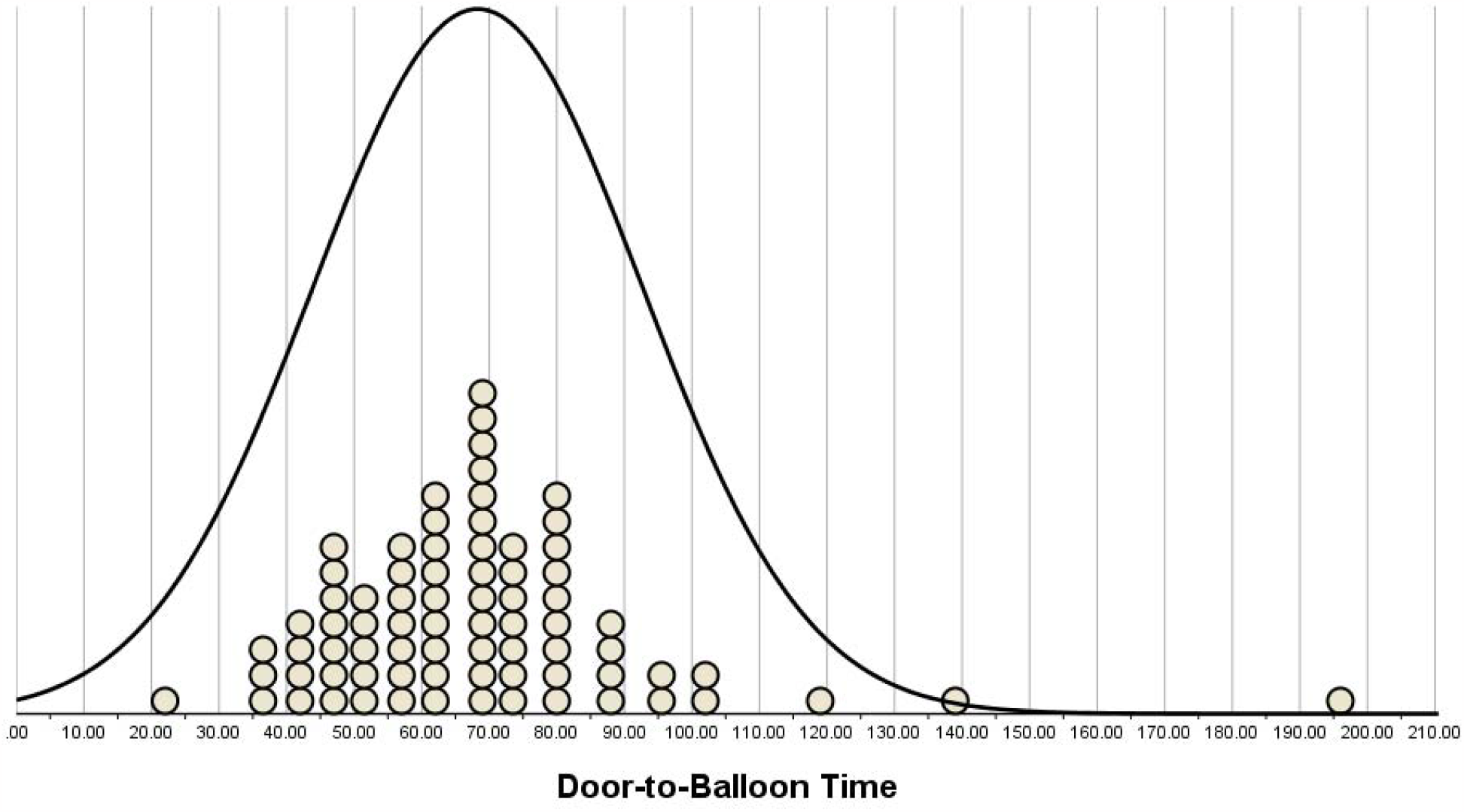
Dot Plot and Normal Distribution Curve of Door-to-Balloon Time in 76 STEMI patients admitted to our hospital during 2011 and 2012.

**Figure 2.**
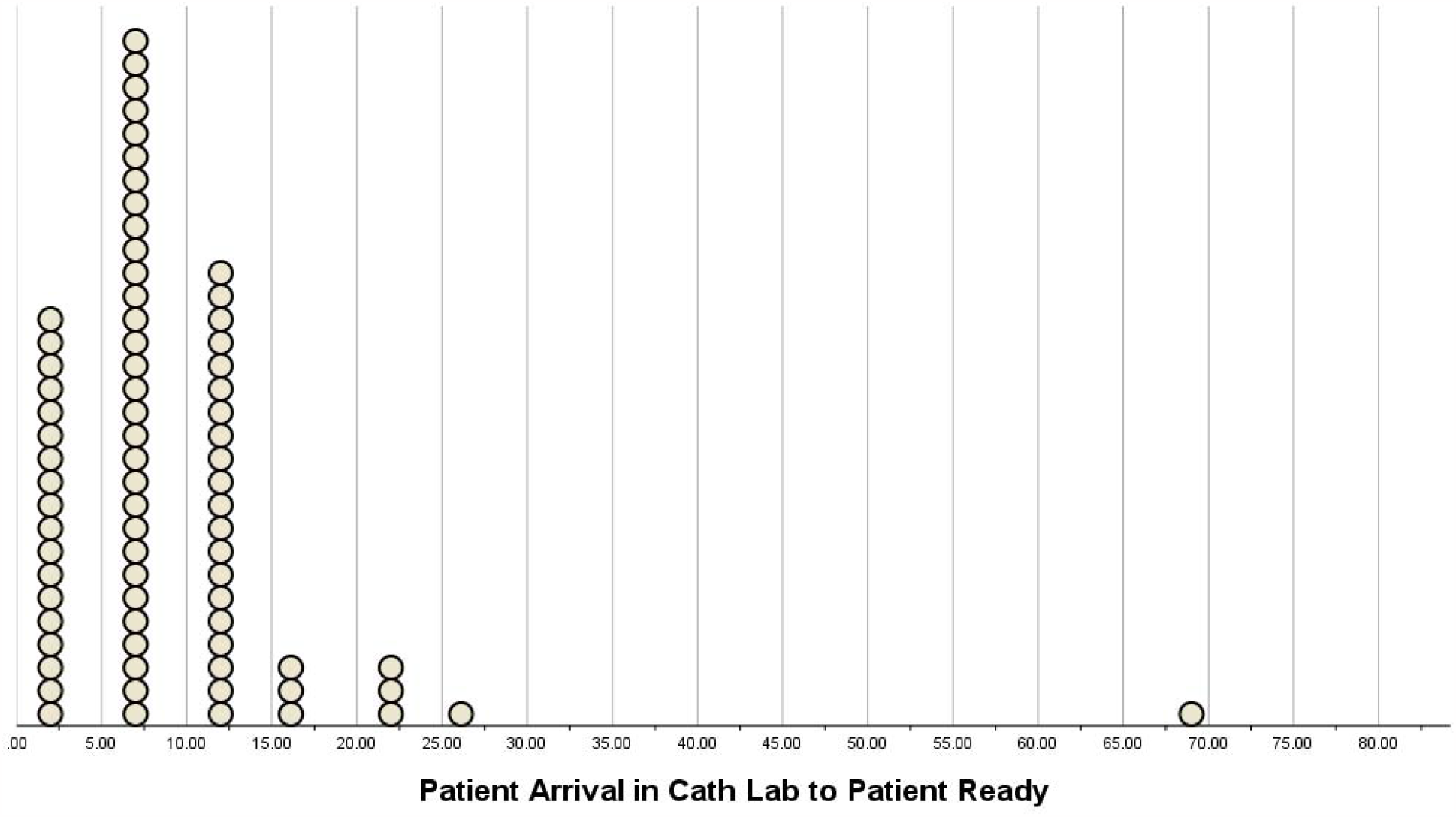
Dot Plot of the time interval of Patient Arrival in Cath Lab to Patient Ready in 76 STEMI patients admitted to our hospital during 2011 and 2012.

**Figure 3.**
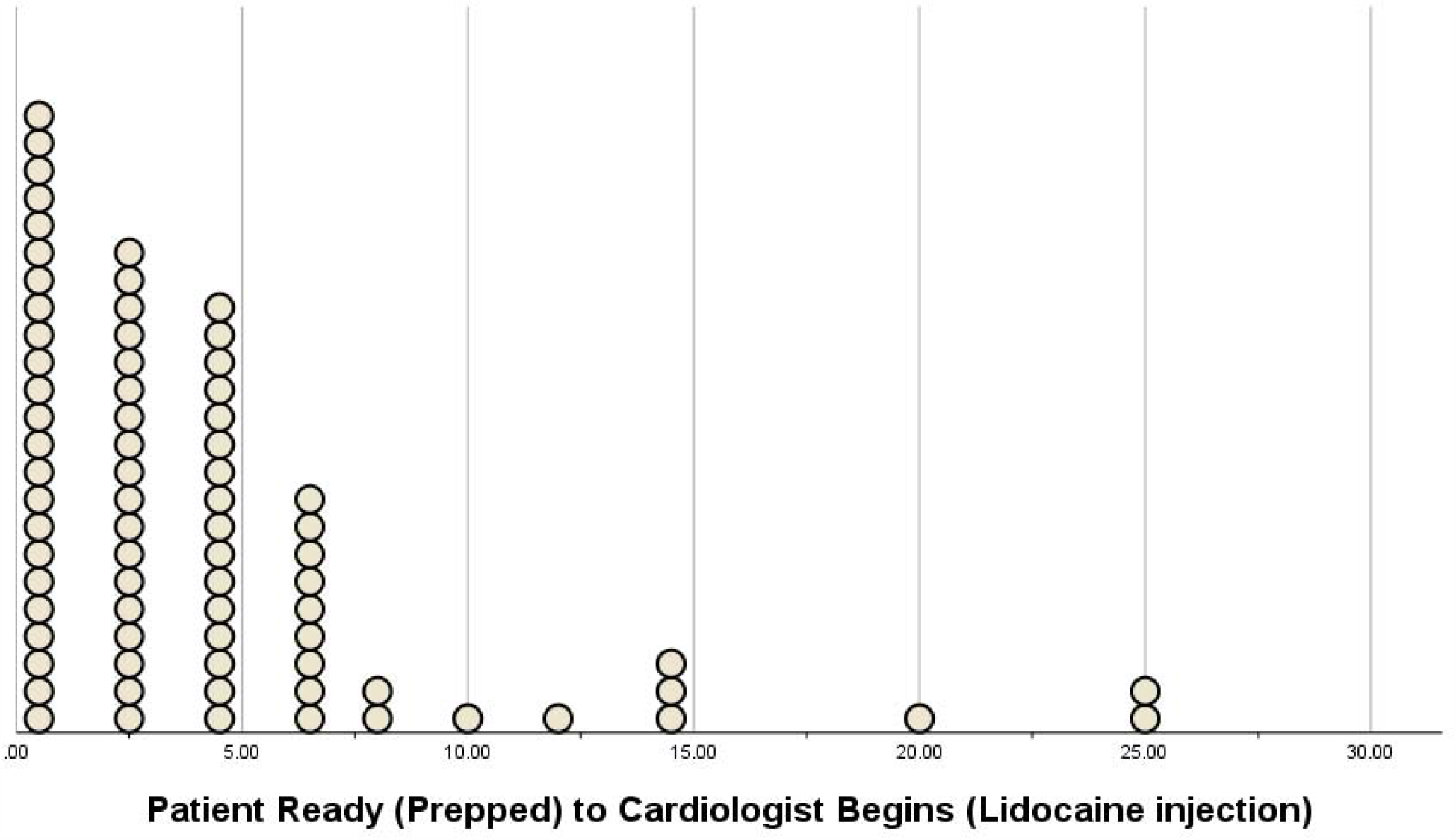
Dot Plot of the time interval of Patient Ready to Cardiologist Begins in 76 STEMI patients admitted to our hospital during 2011 and 2012.

**Figure 4.**
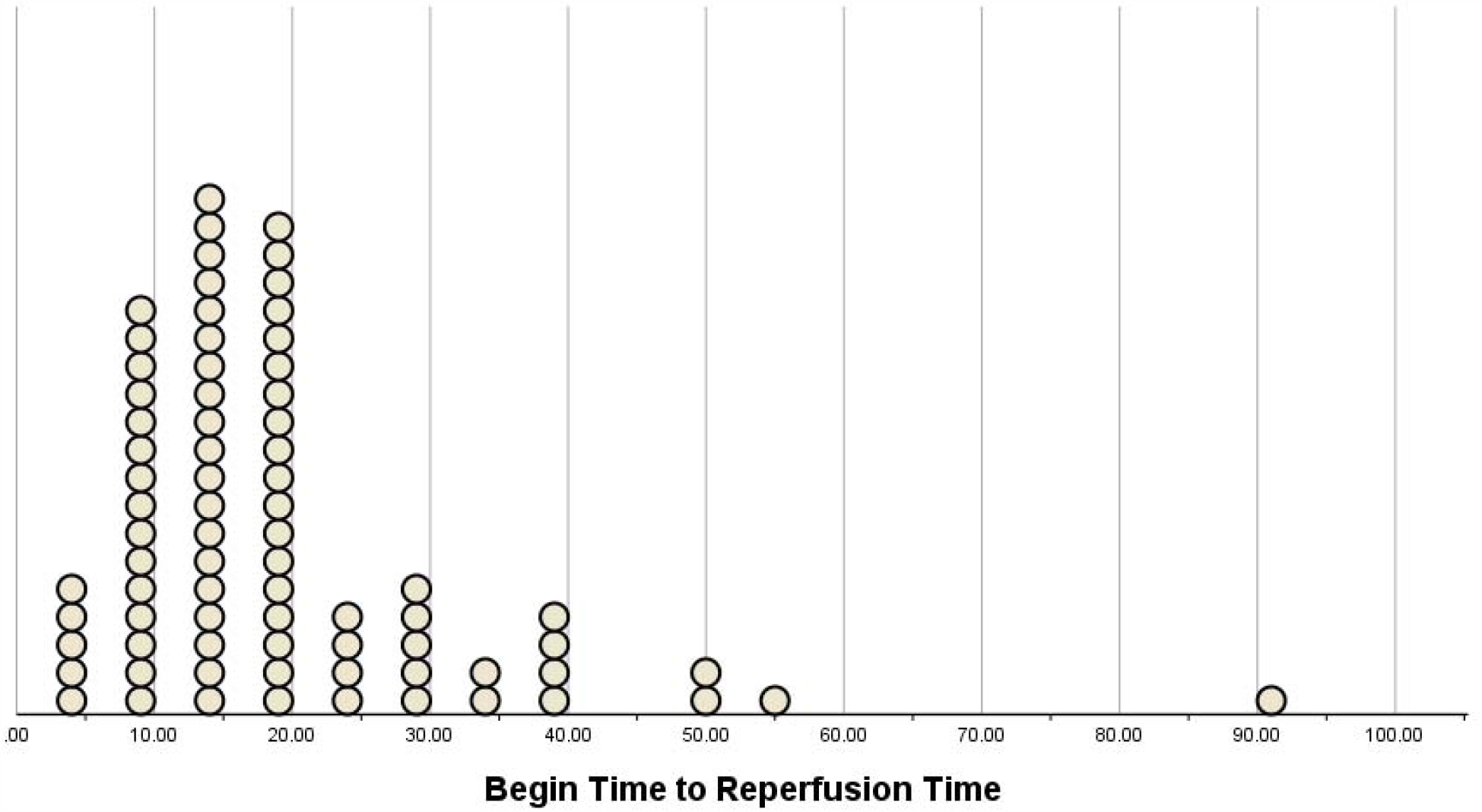
Dot Plot of the time interval of Cardiologist Begins to Reperfusion Time in 76 STEMI patients admitted to our hospital during 2011 and 2012.

The STEMI team arrival to patient arrival in the cardiac catheterization laboratory was significantly higher than the goal of 10 minutes (p-value < 0.001). Patient arrival to ECG, ECG to STEMI read, and STEMI team activation to their arrival compared with the goal of 10, 5, and 25 minutes, respectively, were equal or significantly better (Table 2).

**Table 2.**
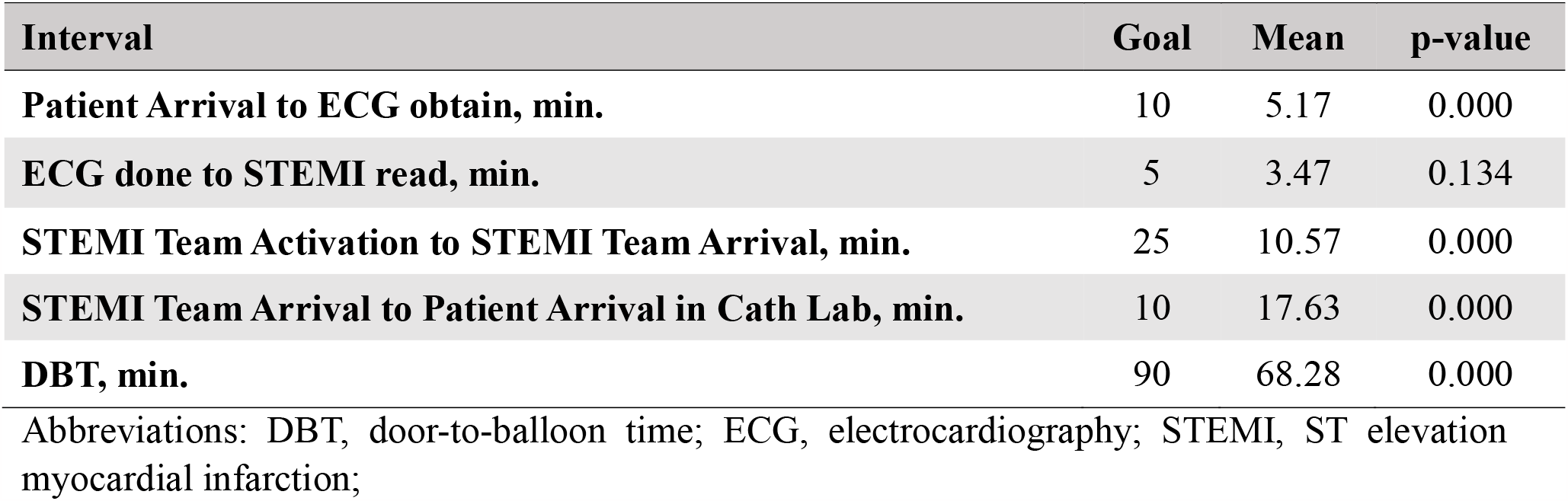
Timeliness Metrics: Goal vs. Actual Performance Analysis.

| **Interval** | **Goal** | **Mean** | **p-value** |
| --- | --- | --- | --- |
| **Patient Arrival to ECG obtain, min.** | 10 | 5.17 | 0.000 |
| **ECG done to STEMI read, min.** | 5 | 3.47 | 0.134 |
| **STEMI Team Activation to STEMI Team Arrival, min.** | 25 | 10.57 | 0.000 |
| **STEMI Team Arrival to Patient Arrival in Cath Lab, min.** | 10 | 17.63 | 0.000 |
| **DBT, min.** | 90 | 68.28 | 0.000 |

Abbreviations: DBT, door-to-balloon time; ECG, electrocardiography; STEMI, ST elevation myocardial infarction;

## Discussion

In our study, the mean DBT was recorded at 68 minutes and 17 seconds. Our findings demonstrated that one of the shortest time intervals was the time for the interventional cardiologist to begin the procedure after the patient was ready (TSP). Our mean TSP time was 4 minutes and 24 seconds with the median of 3 minutes. This result debates that the interventional cardiologists are held accountable for the delay in the DBT for STEMI patients undergoing primary PCI. The most prolonged delay was attributed to the STEMI team and patient arrival in the cardiac catheterization laboratory. This interval was significantly higher than the goal of 10 minutes.

Numerous studies have underscored the pivotal correlation between reperfusion time and the survival rates of acute MI patients. For example, a study by Berger et al., showcased those patients undergoing primary PCI in 60 minutes of ED presentation experienced lower thirty-day mortality compared to those with intervals exceeding 90 minutes (12). Supporting this, an analogous analysis of data from the National Registry of Myocardial Infarction (NRMI) indicated that the reduction in DBT significantly contributed to a progressive decline in in-hospital mortality among all patients treated with primary PCI, dropping from 8.6% in 1994 to 3.3% in 2006 (9). In the research conducted by Zipes et al., it was observed that each half-hour delay in the initiation of early coronary intervention following the onset of acute MI symptoms leads to an 8% increase in the likelihood of mortality over the subsequent year (13). This underscores the criticality of minimizing DBT, even in centers where early coronary intervention is routinely accomplished within 90 minutes (14).

The process of managing STEMI, which encompasses diagnosis and treatment, initiates at the inaugural encounter between medical staff and the patient (15, 16). It’s crucial to emphasize that as per AHA/ACC guidelines, the target duration for a patient’s arrival to obtain an ECG is set at 10 minutes (4). While our mean time of 5 minutes and 10 seconds, closely adheres to this recommendation, we did observe a maximum delay of 27 minutes, signifying an area for potential improvement. Bugami et al. conducted a study with the objective of assessing DBT within an enhancement initiative involving 37 individuals diagnosed with acute STEMI. They also investigated the factors leading to delays in their primary PCI. The findings underscored that the primary factor contributing to prolonged DBT was the delayed identification of STEMI patients (17). Previous studies have emphasized that delays in primary angioplasty are linked to the interval between STEMI diagnosis and staff and patient arrival in the cardiac catheterization laboratory, influenced by consent, personnel availability, and logistical factors. Aspects like dressing, transportation, and equipment availability can also contribute to prolonging this interval (18). Our study underscores this point, highlighting that the most significant delay occurred at the time of the STEMI team arrival to the patient’s arrival in the cardiac catheterization laboratory. The presence of skilled, certified, and extensively trained healthcare personnel constitutes a pivotal factor in the effective management of MI patients (14, 19). It is recommended that an optimal medical team for these cases should consist of an emergency physician, a cardiologist, an interventional cardiologist, a technologist, nursing staff, pharmacists, and laboratory personnel, all working collaboratively to expedite the patient’s transfer to the procedure room. This coordinated effort ensures swift and efficient care delivery (13). In the research conducted by Caputo et al., a concerted effort was made to educate both transport and ED staff about the critical significance of DBT within their institution. Furthermore, an intervention protocol was implemented, which necessitated the presence of cardiac catheterization laboratory staff within a 30-minute window and the interventional staff to perform balloon inflation within the same time frame of the patient being ready. They successfully achieved a noteworthy reduction in DBT (20).

Namdar et al. investigated the impact of implementing a STEMI code on DBT for patients with STEMI. They enrolled 58 STEMI patients, categorizing them into control and intervention groups based on the referral period. Both groups were closely monitored from arrival through emergency services to coronary artery balloon inflation, with times recorded. In the intervention group, a “STEMI code” was activated upon confirmed MI. The study showed a significant reduction in DBT for the intervention group (79.3 ± 27.4 minutes) compared to the control group (113.5 ± 43.6 minutes), with notable significance. This reduction was particularly significant in the cardiac catheterization laboratory to balloon time and overall DBT. The “STEMI code” streamlined care, reducing duplication and optimizing time allocation for acute MIs, subsequently decreasing ED occupancy for more timely admissions (18). Our study, conducted within a STEMI code-designated center, reported a DBT of 68 minutes and 17 seconds, aligning closely with the intervention group’s DBT in their study.

To provide a concise overview, these studies indicate that strategic interventions can lead to notable enhancements in the care of acute MI patients. In our study, patient arrival to ECG obtain, ECG to STEMI read, and STEMI team activation to their arrival compared with the goal of 10, 5, and 25 minutes, respectively, were equal or significantly better. However, there are still modifiable modalities that need to be identified and improved; using pre-hospital ECGs and the ED physician to activate the STEMI code, having a cardiologist on-site 24/7 to be contacted immediately after STEMI code activation, and educating the cardiac catheterization laboratory personnel on the importance of being present in the procedure room lab in less than 20 minutes of STEMI code announcement as well as contact an interventional cardiologist promptly once the diagnosis is confirmed are among the most important factors (21). Each hospital staff should be alert about these modifications and improve the shortcomings of the factors that lengthen the DBT in their hospital, leading to better prognosis and lower mortality of STEMI patients. It is crucial to underscore the pivotal role played by the time lapse between symptom onset and hospital arrival, a factor of paramount importance in the setting of STEMI (22). Elevating the prognosis and overall outcomes for STEMI patients hinges on the diligent focus and proactive measures applied during this critical phase as well as DBT. We have no clear explanation why the cath lab team arrival to the patient arrival time in the cardiac catheterization laboratory is higher than national expectations. This might be related to the delay in the patient transport time between the ER and the cath lab. As value-based cardiologist compensation is gaining popularity, we should avoid using DBT as a cardiologist metric as based on our findings, cardiologists play the smallest role in the DBT. (23)

## Limitations

While this study delves into the intricate nuances of various time frames encompassing the total DBT and proposes potential solutions for enhancing the outcomes of STEMI patients, it is crucial to recognize that these timeframes may vary significantly across different medical centers. It is imperative to acknowledge the constraints of this research, which is primarily attributed to its single-center focus. The limited external validity of our research necessitates future multi-center studies to corroborate and generalize our proposed solutions for enhancing outcomes in STEMI patients. Furthermore, it is recommended that upcoming research includes diligent patient follow-up to assess both DBT and patient prognosis. Additionally, a concerted effort should be made to prioritize and expedite the time from symptom onset to hospital admission. We do not have time separation based on STEMI occurring during day or after hours that could have been useful to detect any DBT time delay based on the prestation time.

## Conclusions

Our data is the first to evaluate delays related to DBT revealing the least delay occurring due to the late arrival of Interventional cardiologists. Our data emphasizes the importance of performing a detailed time analysis of the DBT delay in order to objectively determine the actual areas of delay and provide a future pathway to improve them since we have specifically detected a delay in STEMI team and patient arrival to the catheterization laboratory as the main delay in the DBT time.

## Data Availability

not available

